# Efficacy of the glucagon-like peptide-1 agonist Exenatide in patients undergoing coronary artery bypass grafting or aortic valve replacement – a randomized double-blind clinical trial

**DOI:** 10.1101/2024.10.15.24315567

**Authors:** Jesper Kjaergaard, Christian Holdflod Møller, Sebastian Wiberg, Astrid Duus Mikkelsen, Hasse-Møller Sørensen, Hanne Ravn, Jesper Ravn, Peter Skov Olsen, Dan Høfsten, Søren Boesgaard, Lars Køber, Jens Christian Nilsson, Christian Hassager

## Abstract

**Importance:** Glucagon-like peptide-1 (GLP-1) agonists have been proven beneficial in reducing risk of and injury associated with several cardiovascular diseases. The efficacy in cardiopulmonary bypass (CPB)-assisted cardiac surgery is unknown.

**Objective:** This trial aimed to investigate the efficacy of an infusion of the GLP-1 antagonist Exenatide during and after open heart surgery in reducing risk of death and major organ failure.

**Design:** Randomized, double-blinded, 2-by-2 factorial design, clinical trial, also including liberal (FiO2 of 100%) or restrictive (FiO2 of 50%) oxygenation during and after bypass. The present paper presents the results of the Exenatide intervention.

**Setting:** Single site, tertiary heart center.

**Participants:** Adult patients undergoing elective cardiopulmonary bypass-assisted coronary artery bypass grafting and/or aortic valve replacement.

**Intervention:** Infusion of 17.4 µg og Exenatide or placebo during cardiopulmonary bypass and the first hour after weaning thereof

**Main outcomes:** The main outcome was time to a composite endpoint consisting of death, stroke, renal failure requiring dialysis, or new/worsening heart failure during follow-up. Secondary endpoints included occurrence of prespecified adverse events.

**Results:** A total of 1389 patients were included in the analyses. Within a follow-up period of median of 5.9 years (min – max; 2.5 – 8.3 years), 170 patients (24%) in the Exenatide group and 165 patients (24%) experienced a primary endpoint. We found no difference in time to first event between patients randomized to FiO_2_ 50% versus FiO_2_ 100% (HR 1.0 [95%CI 0.83 – 1.3], *p* = 0.80). We found no significant difference in rates of adverse events between the two groups.

**Conclusions and Relevance:** Exenatide during cardiopulmonary bypass and weaning thereof did not significantly reduce the incidence of death, stroke, renal failure, or new/worsening heart failure in patients undergoing coronary artery bypass grafting and/or aortic valve replacement.

**Trial registration:**

1. Danish Medicines Agency: Protocol no. HJE-PHARMA-001, EudraCT no. 2015-003050-41, 2^nd^ of October 2015
2. Local Ethics Committee “Videnskabsetisk komité C, Region Hovedstaden”: No. H-15010562

www.clinicaltrials.gov: ID no. NCT0267393

**Key Points:** *Question:* Is a single dose of Exenatide effectively reducing risk of death or major organ injury in patients undergoing cardiopulmonary bypass (CPB)-assisted cardiac surgery?

*Findings:* 1400 patients undergoing coronary artery bypass grafting and/or aortic valve replacement were randomized to 17,4 µg of Exenatide or placebo during CPB and the first hour after weaning. The hazard ratio (95%CI) for time to the first occurring composite endpoint consisting of death, stroke, renal failure requiring dialysis, and new/worsening heart failure was 1.0 (0.83 – 1.3).

*Meaning:* Exenatide infusion during CPB-assisted cardiac surgery does not improve outcomes.

## Introduction

Cardiopulmonary bypass (CPB) assisted cardiac surgery is associated with a risk of organ injury that may result in permanent sequelae or death. The precipitating indication for surgery, for example coronary artery disease or aortic valve disease caused by atherosclerosis, combined with perioperative hemodynamic disturbances increase the risk of ischemia- and reperfusion-induced organ injury. Furthermore, the application of CPB result in a systemic inflammatory response and hemolysis, which may cause or aggravate organ damage^1^. Within the first year from surgery, the risk of major cardiovascular events (MACE) and mortality is 6-7% and 3-4%, respectively^2,3^, however, in patients older than 70 years, the 30-day MACE rate increases to 10%^4^. Currently, no specific pharmacological interventions have been shown to reduce the risk of organ injury or adverse events during on-pump cardiac surgery.

Glucagon-like peptide-1 (GLP-1) analogs stimulate glucose-dependent insulin release, and several GLP-1 analogs are approved for treatment of type II diabetes, and obesity. In pre-clinical models of both stroke and acute myocardial infarction (MI), GLP-1 analogs have been shown to reduce the final infarct size^5–10^. In humans with MI, GLP-1 analogs administered before revascularization have been shown to increase myocardial salvage, and to result in a smaller final infarct size in a subset of patients with a limited ischemia time^11–14^. In comatose patients, resuscitated after out-of-hospital cardiac arrest, administration of the GLP-1 analog exenatide improved lactate clearance after admission^15^. Finally, exenatide reduced reperfusion injury in patients with myocardial infarction^16^. Notably, exenatide did not increase the rate of adverse events^15,17^.

The aim of the present study was to assess, whether an infusion with the GLP-1 analog exenatide administered prior to and during CPB-assisted coronary artery bypass grafting and/or aortic valve replacement would reduce mortality and severe organ injury, defined as stroke, renal failure or heart failure.

## Methods

The GLORIOUS trial was a single center, investigator-initiated, 2-by-2 factorial design, randomized controlled trial. Patients undergoing elective or subacute Coronary Artery Bypass Grafting (CABG) and/or Aortic Valve Replacement (AVR) were randomized to treatment with Exenatide versus placebo (in a double-blinded intervention) and to receive FiO2 of 50% or 100% during CPB and the first hour after weaning from CPB (blinded for outcome assessors and participants). Patients were randomized in a tertiary heart center in Denmark via an internet-based randomization algorithm using permuted blocks of 4, 8, or 12 participants. The randomization was stratified by AVR. The results of the oxygenation strategy will be reported separately. The study protocol is available on clinicaltrials.gov (NCT02673931) and the design paper has been published^18^.

The Danish Medicines Agency and the Regional Ethics Committee of the Capital Region of Denmark approved the trial protocol prior to trial initiation. The trial was designed and conducted by the steering committee. Authors JK, SCW and ADM analyzed the collected data, and the first author wrote the first manuscript draft. All authors vouch for the accuracy and completeness of the data, analyses, and final article.

### Patients

We included adult patients (≥18 years of age) undergoing elective or subacute CABG and/or AVR irrespective of other concomitant valve surgery. Key exclusion criteria included acute surgery, active treatment with GLP-1 analogs, and known allergy to study drugs. A comprehensive list of exclusion criteria ca be found in the trial protocol^18^, and in supplementary appendix 1.

### Endpoints

The primary endpoint was time to the first occurring of the following primary endpoint during follow-up:

1. Death from any cause, or
2. The occurrence of any of the following adverse events, adjudicated by an endpoint committee blinded for treatment allocation:

a. Renal failure requiring any type of renal replacement therapy.
b. Stroke, defined as any sign or symptom of neurological dysfunction persisting for more than 24 hours, determined by the treating physician based on clinical information including CT-scan.
c. New onset or worsening heart failure defined as need for mechanical circulatory support at the ICU, inability to close the sternum due to hemodynamic instability and/or need for inotropic hemodynamic support more than 48 hours after initiation of the first surgical procedure after randomization. In addition, any

admission for heart failure during follow-up after discharge from the index admission.

Secondary endpoints included time in days to occurrence of each component of the primary endpoint, and the incidence of the following safety endpoints: Surgical site infection, doubling of S-creatinine or urine output below 0.5ml/kg/hour for minimum 12 hours (KDIGO stage 2 or higher^19^), hypoglycemia (blood glucose < 3mmol/L), pancreatitis, a reduction of LVEF of 50% or more compared to baseline, re-operation for any cause, post-surgery MI (Type 5 MI), and re-admission for cardiovascular causes^18^.

### Intervention

Patients were randomized in a one-to-one ratio to a 6 hour and 15 minutes infusion of either 17.4 μg of exenatide (Byetta®, Lilly) or placebo initiated at time of anesthesia prior to surgery. Both the active treatment and placebo infusions consisted of 248.5 mL of isotonic (0.9%) NaCl with 1.5 mL of 20% human albumin. A total of 25 μg of exenatide was added to the active treatment infusions kits. A total volume of 174 mL was administered over the 6 hour and 15-minute infusion. The infusion kit was prepared in a neighboring ward by trained nurses, who had otherwise no involvement in the trial. Thereafter the infusion kit was transported to the operating theatre, maintaining blinding for the allocation of study drug to all patients, clinical staff, and all trial staff.

### Treatment protocol

All participants received standard care. Anesthesia was conducted in line with guidelines^20^ with the standard use of fentanyl, propofol and rocuronium for induction followed by anesthesia with sevoflurane and remifentanil infusion. Peripheral veins were canulated and invasive arterial blood pressure monitoring was applied prior to induction of anesthesia. After induction of anesthesia, a central venous catheter was placed. Transesophageal echocardiography and pulmonary artery catheter were used at the discretion of the clinical staff. During CPB, a PaCO_2_ from 4.5 to 6.0 kPa, a hematocrit above or equal to 21%, a body temperature above or equal to 36.5°C were targeted. A fixed pump of 2.4 L/min/m^2^ body surface area plus 10-20% was generally applied.

### Sample Size

No interaction between the two study interventions was expected *a priori*, and the sample size estimation did not account for interactions. If 1400 patients were included, the trial would be able to show a relative reduction in the primary endpoint of 25% from an expected event rate of 23% (i.e. 323 events; based on data from the surgical register at the trial site) with a power of 80% at an alpha-level of 0.05 (two-sided). The trial was designed as event driven, with inclusion of 1400 patients being followed until a total of 323 events had been reached.

### Statistical analyses

All analyses were conducted on the intention-to-treat population with a two-sided significance level of 0.05 being applied. Categorical variables will be presented as numbers (percentages), and differences between groups are tested with the chi-square test or Fisher’s exact test as appropriate. Continuous variables are presented as mean ± SD if normally distributed or otherwise as median (interquartile range), and differences between groups will be tested with the independent sample t-test or the Wilcoxon’s rank sum test as appropriate. For the primary endpoint analysis, Kaplan-Meier curves will be displayed and compared using the two-sided log-rank test for testing of differences between strata. A Cox proportional hazard model will be applied with adjustment for age, sex, body mass index, year of inclusion, procedure (CABG vs AVR vs CABG+AVR), known alcohol or drug abuse, known heart failure, known dialysis, known pulmonary disease, known diabetes, previous stroke, previous myocardial infarction, previous PCI, previous CABG, previous AVR, and length of cardiopulmonary bypass. As an exploratory analysis collinearity between exenatide and the oxygenation treatment strategy will be explored. As sensitivity analyses, the primary analyses will be repeated with a follow-up time limited to 180 days (supplementary appendices).

## Results

From February 2016 through December 2021, a total of 1400 patients were randomized in the trial, 11 of whom withdrew their consent and the intention-to-treat population consisted of 1389 patients (Figure 1). The analyses were conducted on this population as intention-to-treat. Two (0.14%) patients in the placebo group died between randomization and the surgical procedure, and in 17 (1.2%) patients, the planned surgical procedure was changed or cancelled after randomization (Figure 1). Final follow-up was completed in June 2024.

**Figure 1.**
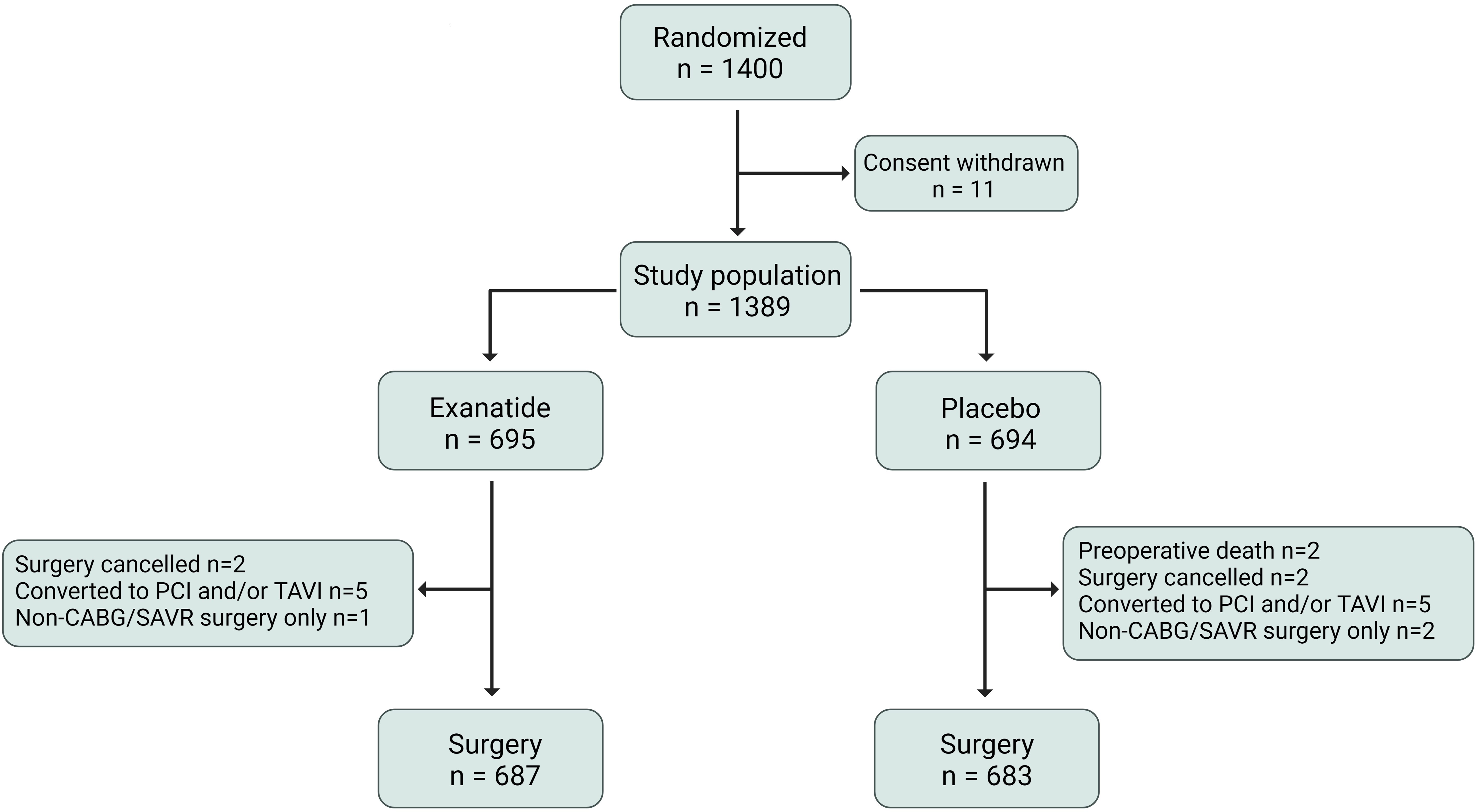
Consort diagram. PCI: percutaneous coronary intervention; TAVI: transcatheter aortic valve implantation; CABG: coronary artery bypass grafting; SAVR: surgical aortic valve replacement

### Patient characteristics

Patients were 68 years of age (IQR 60 – 74 years) with 17% being female in the two randomization groups. Baseline characteristics between intervention groups appeared well balanced (Table 1). A total of 206 (15%) patients had type 2 diabetes. The majority of patients (68%) underwent isolated CABG. Surgical characteristics between the intervention groups appeared well balanced (Table 2).

**Table 1.** Baseline characteristics stratified by treatment allocation.

|  |  | **Exenatide**<br>**(n=695)** | **Placebo**<br>**(n=694)** |
| --- | --- | --- | --- |
| Age – years | Median [IQR] | 68 [61, 74] | 68 [60, 73] |
| Female sex | n(%) | 119 (17) | 114 (16) |
| Caucasian | n(%) | 673 (98) | 685 (99) |
| Body mass index – kg/m<sup>2</sup> | Median [IQR] | 27.1 [25, 30] | 27.1 [25, 30] |
| **Comorbidities and performance** |  |  |  |
| Smoking, current or prior | n(%) | 441 (64) | 450 (65) |
| Alcohol consumption | N(%) | 5 (0.7) | 9 (1.3) |
| Heart failure | n(%) | 113 (16) | 112 (16) |
| Hypertension | n(%) | 437 (63) | 461 (67) |
| Type 2 diabetes | n(%) | 104 (15) | 102 (15) |
| Type 1 diabetes | n(%) | 8 (1.2) | 13 (1.9) |
| Pulmonary disease | n(%) | 78 (11) | 75 (11) |
| Ischemic heart disease | n(%) | 204 (30) | 206 (30) |
| Previous myocardial infarction | n(%) | 127 (18) | 136 (20) |
| Previous percutaneous coronary int. | n(%) | 101 (15) | 94 (14) |
| Previous coronary artery bypass graft | n(%) | 7 (1.0) | 4 (0.6) |
| Previous valve surgery | n(%) | 4 (0.6) | 14 (2.0) |
| Previous stroke | n(%) | 65 (9.4) | 59 (8.5) |
| Pacemaker or ICD | n(%) | 12 (1.7) | 20 (2.9) |
| Dialysis | n(%) | 12 (1.7) | 11 (1.6) |
| Frailty score ≥4 | n(%) | 172 (34) | 182 (37) |
| NYHA class ≥3 | n(%) | 145 (27) | 126 (24) |
| CCS class ≥3 | n(%) | 72 (14) | 58 (11) |
| NT-proBNP – pmol/L | median [IQR] | 36.6 [ 15, 100] | 35.3 [14, 97] |
| Creatinine – µmol/L | median [IQR] | 84 [74, 98] | 86 [ 76, 100] |
| **Medications** |  |  |  |
| Acetylsalicylic acid | n(%) | 440 (64) | 444 (64) |
| Non-vitamin K oral anticoagulants | n(%) | 52 (7.6) | 64 (9.3) |
| Angiotensin-converting enzyme inhibitor | n(%) | 189 (28) | 161 (23) |
| Angiotensin receptor blocker | n(%) | 170 (25) | 193 (28) |
| Statins | n(%) | 495 (72) | 505 (73) |
| Beta blockers | n(%) | 296 (43) | 322 (47) |
| Thiazides | n(%) | 113 (16) | 108 (16) |
| Insulin | n(%) | 35 (5.1) | 31 (4.5) |

ICD: implantable cardiac defibrillator; NYHA: New York Heart Association; CCS: Canadian cardiovascular society; NT-proBNP: N-terminal pro brain natriuretic peptide

**Table 2.** Perioperative characteristics stratified by treatment allocation.

|  |  | **Exenatide**<br>**(n=695)** | **Placebo**<br>**(n=694)** |
| --- | --- | --- | --- |
| Primary indication for surgery |  |  |  |
| CABG | n(%) | 360 (68) | 358 (69) |
| AVR | n(%) | 166 (32) | 161 (31) |
| Performed surgery: |  |  |  |
| CABG | n(%) | 469 (68) | 468 (68) |
| AVR | n(%) | 151 (21) | 153 (22) |
| CABG+AVR | n(%) | 67 (9.7) | 62 (9.0) |
| For CABG: No. of diseased coronary vessels |  |  |  |
| 1 | n(%) | 38 (7.5) | 40 (8.0) |
| 2 | n(%) | 114 (23) | 95 (19) |
| 3 | n(%) | 235 (46) | 245 (49) |
| For AVR: Prothesis type |  |  |  |
| Biological | n(%) | 154 (23) | 156 (24) |
| Mechanical | n(%) | 50 (7.5) | 53 (8.0) |
| Additional surgery: |  |  |  |
| Ascending aorta | n(%) | 4 (0.6) | 9 (1.4) |
| Mitral valve | n(%) | 4 (0.6) | 3 (0.4) |
| Tricuspid valve | n(%) | 1 (0.2) | 1 (0.2) |
| Patent foramen ovale closure | n(%) | 1 (0.2) | 0 |
| Left atrial appendix closure | n(%) | 1 (0.2) | 0 |
| Duration of: |  |  |  |
| Cardiopulmonary bypass – min. | median [IQR] | 89 [ 72, 118] | 86 [ 69, 111] |
| Aortic cross clamp – min. | median [IQR] | 57 [42, 78] | 54 [40, 77] |
| Reperfusion – min. | median [IQR] | 23 [17, 30] | 22 [17, 29] |

CABG: coronary artery bypass grafting; AVR: aortic valve replacement

### Blood glucose levels

In the operating room, prior to induction of anesthesia and prior to intervention, median blood glucose levels were similar between the two intervention groups (Figure 2). After initiation of the intervention, patients randomized to exenatide had significantly lower blood glucose levels after initiation of the exenatide infusion (Figure 2). However, rates of hypoglycemia below 3 mmol/L were rare and non-significantly different between intervention groups (Table 3).

**Figure 2.**
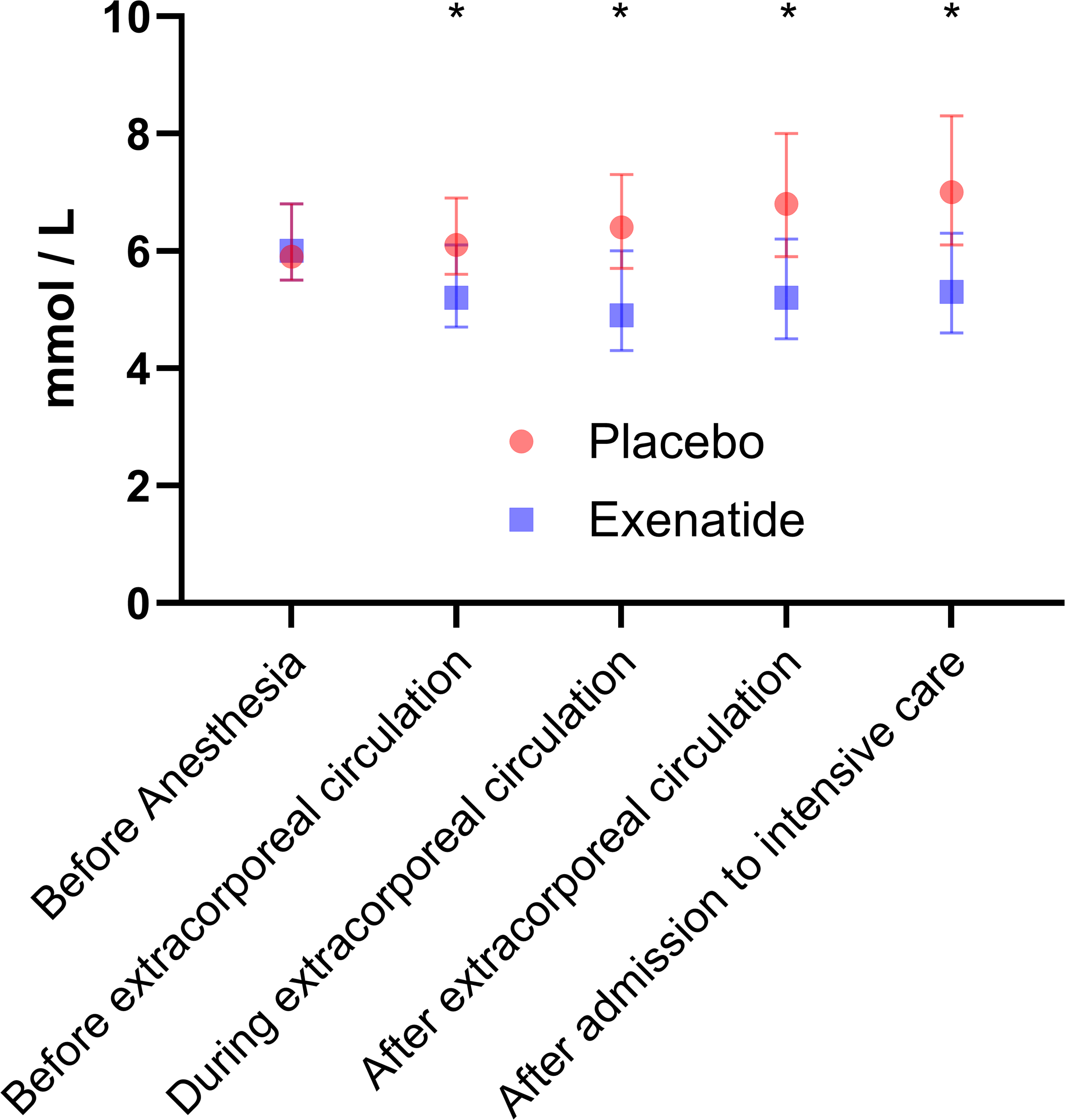
Blood glucose levels during surgery stratified by treatment allocation. Levels of blood glucose from prior to induction of anesthesia (i.e. prior to intervention) to admission to the intensive care unit. * denotes p < 0.001 (Wilcoxon’s Rank Sum test)

**Table 3.** Safety events stratified by treatment allocation.

|  | **Exenatide**<br>**n=695** | **Placebo**<br>**n=694** | **p** |
| --- | --- | --- | --- |
| Death | 97 (14) | 87 (13) | 0.48 |
| Surgical site infection | 19 (2.7) | 14 (2.0) | 0.38 |
| Acute kidney injury | 33 (4.8) | 37 (5.3) | 0.62 |
| Hypoglycemia within 12 hours | 2 (0.29) | 0 (0.0) | 0.50 |
| Pancreatitis | 2 (0.3) | 1 (0.1) | 0.99 |
| Reduction of LVEF > 50% from baseline | 10 (1.4) | 8 (1.2) | 0.64 |
| Re-operation for any cause | 16 (2.3) | 21 (3.0) | 0.40 |
| Post-surgery myocardial infarction | 10 (1.4) | 4 (0.58) | 0.11 |
| Re-admission for cardiovascular causes | 185 (27) | 178 (26) | 0.68 |

LVEF: Left ventricular ejection fraction
Acute kidney injury defined as doubling of S-creatinine of urine output below 0.5ml/kg/min for over 12 hours. Hypoglycemia defined as blood glucose below 3 mmol/L. Pancreatitis defined as serum amylase above 3 times upper normal limit.

### Endpoints

The primary endpoint occurred in 170 (24%) patients in the Exenatide group and 165 (24%) patients in the placebo group, with no significant difference in time to the first primary endpoint between allocation groups (unadjusted HR 1.0 [95%CI 0.83 – 1.3], P_logrank_ = 0.80, Figure 3). This did not change in the pre-defined cox proportional hazard model after adjustment for potential confounding factors (HR 0.99, 95% confidence interval 0.78-1.3, *p* = 0.93). We found no significant interaction between exenatide and the oxygen intervention (*p*_interaction_=0.40) Furthermore, we found no significant differences in time to death, in time to stroke, in time to renal failure requiring dialysis, or in time to worsening or new onset heart failure considering competing risk of death (Figure 4). We found no significant differences in pre-defined adverse events between allocation groups (Table 3).

**Figure 3.**
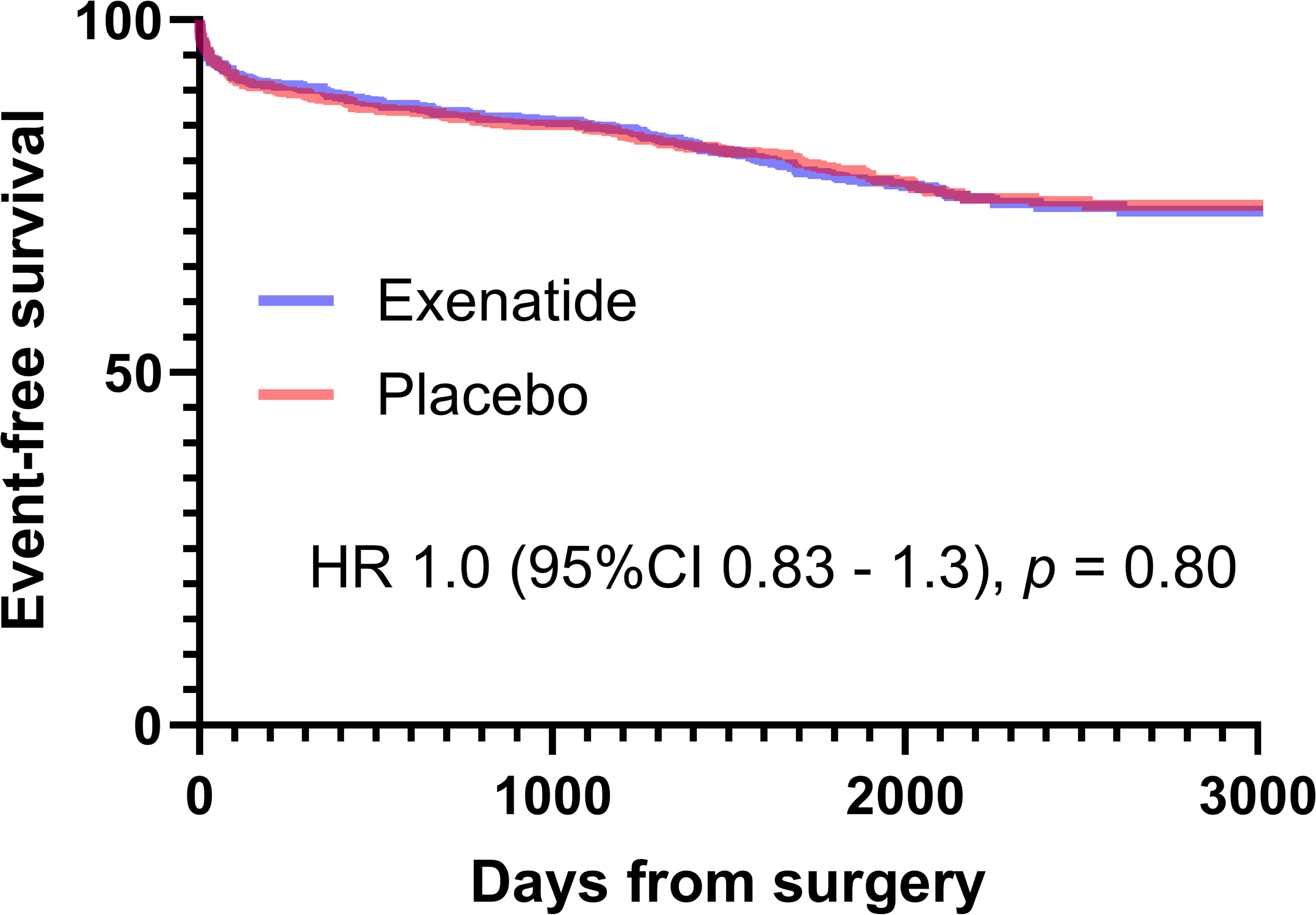
Time to first primary endpoint between the two treatment allocations.

**Figure 4.**
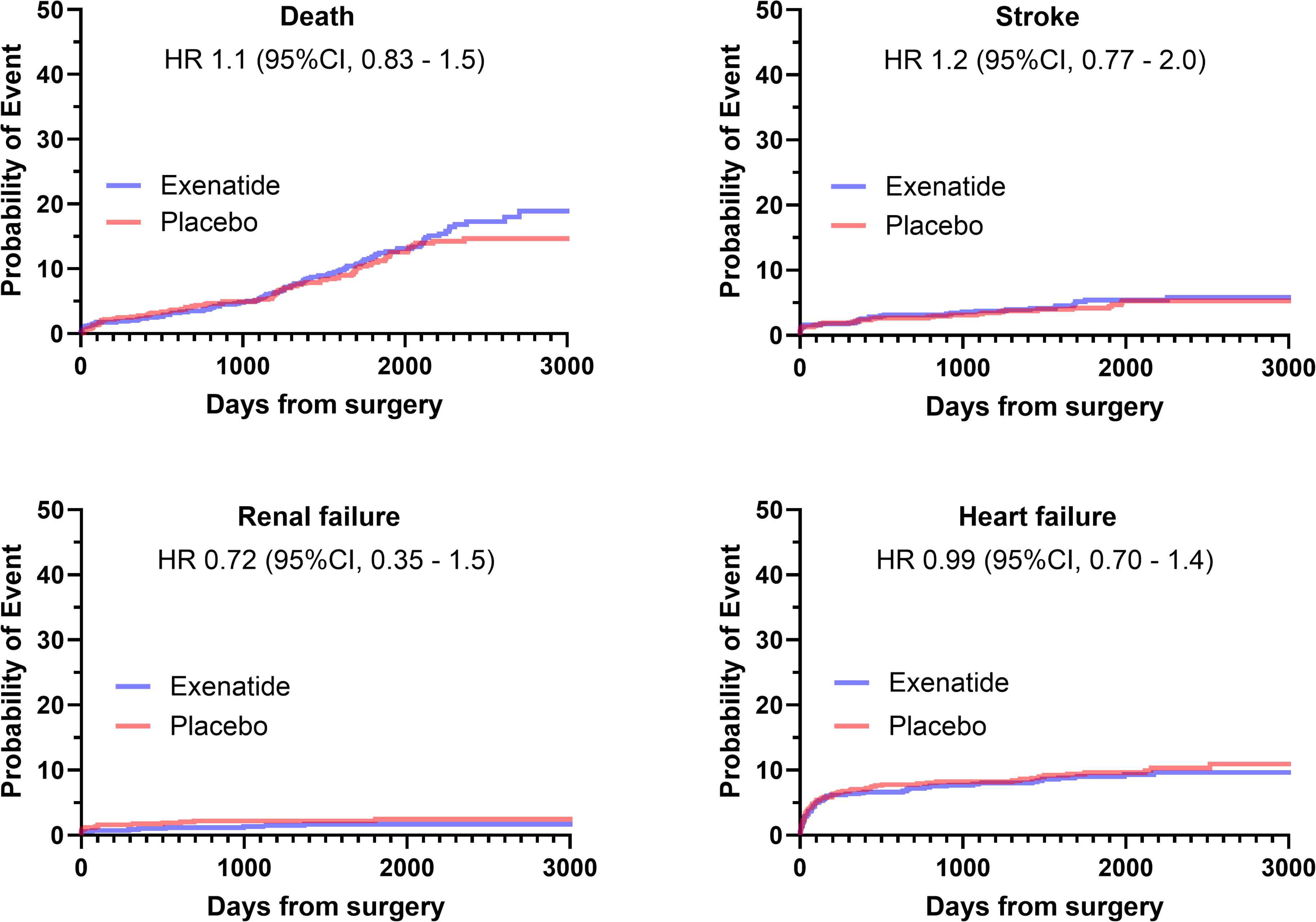
Time to the individual components of the co-primary endpoint stratified by treatment allocation. Models of stroke, renal failure, and now onset or worsening heart failure take competing risk of death into account. Stroke is defined as clinical stroke as diagnosed by the clinical physician. Renal failure is defined renal failure requiring renal replacement therapy. Heart failure is defined as need for mechanical circulatory support at the ICU, inability to close the sternum due to hemodynamic instability and/or need for inotropic hemodynamic support more than 48 hours after initiation of the first surgical procedure after randomization. In addition, any admission for heart failure during follow-up after discharge from the index admission.

In pre-planned sensitivity analyses, we found no difference in time to the first occurring co-primary endpoint within 180 days between the two allocation groups (P_logrank_ = 0.78, Supplementary Appendix 2). Furthermore, we found no significant differences in time to the individual co-primary endpoints within 180 day (Supplementary Appendix 3). The efficacy of Exenatide in selected subgroups did not show significant differential effects in patients with or without preexisting diabetes, heart failure or renal disease, but, patients with previous stroke may have benefit of Exenatide (Supplementary appendix 4).

## Discussion

The present study found no significant difference in a composite endpoint of death, stroke, heart failure and renal failure requiring dialysis in patients undergoing CPB-assisted cardiac surgery treated with a perioperative infusion of a GLP-1 analog versus placebo. To our knowledge, this is the first trial adequately powered towards a clinical endpoint to investigate if perioperative infusion of Exenatide prevent organ injury.

GLP-1 is an incretin hormone stimulating insulin production in response to food intake^21^, and GLP-1 analogs have accordingly been approved for treatment of type 2 diabetes. GLP-1 analogs have been suggested to have organ protective effects during ischemia and reperfusion through complex intracellular pathways ^22–24^.

In an outpatient setting, several previous trials in humans have investigated the effects of GLP-1 analogs on MACE in patients with type 2 diabetes^25^. The EXSCEL trial randomized 14,752 patients with type 2 diabetes to exenatide versus placebo and showed a borderline difference in MACE after a median follow-up of 3.2 years (HR 0.91 [0.83-1.0, p=0.06])^26^. The LEADER trial randomized 9,340 patients with type 2 diabetes and a high cardiovascular risk to treatment with the GLP-1 analog liraglutide versus placebo and found a significant reduction in MACE (HR 0.87 [0.78-0.97], p=0.01) and all-cause mortality (HR 0.85[0.74-0.97], p=0.02) after a median follow-up of 3.8 years^27^. Furthermore, liraglutide has a significant positive effect on a secondary composite outcome on renal function^27^. The REWIND trial randomized 9901 patients with type 2 diabetes, a previous cardiovascular event or cardiovascular risk factors to the GLP-1 analog dulaglutide and found significant reduction in MACE (HR 0.88 [0.79-0.99], p=0.026^28^. The Harmony Outcomes Trial randomized 9,463 type 2 diabetes patients with known cardiovascular disease to the GLP-1 analog albiglutide versus placebo and found a significant reduction in MACE (HR 0.78 [0.68-0.90], p=0.0006) after a median of 1.6 years^29^. The SUSTAIN-6 trial randomized 3,297 type 2 diabetes patients to the GLP-1 analog Semaglutide versus placebo and found a significant reduction in MACE (HR 0.74 [0.58-0.95], p < 0.001 ^29^. As such, there is solid evidence showing that GLP-1 analogs reduce the risk of MACE in patients with type 2 diabetes^30^. In addition, the SELECT trial randomized 17,604 patients without diabetes but with preexisting cardiovascular disease and a BMI equal to or greater than 27 to Semaglutide versus placebo and found a significant reduction in cardiovascular events (HR 0.8 [0.72-0.90], p<0.001)^31^.

In the perioperative setting, smaller trials have investigated the effects of GLP-1 analogs. A meta-analysis from 2018 investigated the effects of GLP-1 analogs versus placebo on glycemic control and the prevalence of hypoglycemia in the operating room or critical care setting. The analysis found that GLP-1 analogs overall resulted in improved glycemic control without a significant increase in hypoglycemia^32^. This is in accordance with the results of the present trial suggesting that GLP-1 analogs may be an attractive alternative to perioperative insulin treatment. Pre-operatively administered GLP-1 analog may delay gastric emptying and be associated with an increased risk of aspiration prior to intubation, and it remains debated whether GLP-1 analogs prescribed for treatment of diabetes should be continued through the perioperative period^33,34^. We did not find any difference in a composite clinical endpoint including heart failure, however the effect of GLP-1 specifically cardiac function after cardiac surgery remains to be investigated. The subgroup analysis showed a benefit in patients with previous strike, which should be interpreted with caution until confirmed in other populations.

The presented results should be interpreted with some caution due to the following limitations. The mechanisms behind the suggested organ protective effects of GLP-1 analogs remain poorly understood, and data on optimal dose and type of GLP-1 analog for the perioperative setting remain scarce. The short duration of exposure to the drug has previously been effective in reducing myocardial injury in myocardial infarction ^16^, but a different effect with linger exposure to the study drug cannot be ruled out by the present design. The trial event rate was lower than expected, and accordingly, the follow-up period was prolonged. However, sensitivity analyses did not suggest any treatment effects in the perioperative or short-term follow-up either. The single-center trial design may limit generalizability to other settings and the study population mainly consisted of low-risk patients undergoing elective surgery and the results should not be applied in higher risk populations.

Several meta-analyses have been published, comparing different GLP-1 analogues and the overall beneficial effect of the compounds seem to be a class effect^35,36^, however Lixisenatide may not be equally efficacious in preventing major cardiovascular outcomes as the remainder of the GLP-1 analogues^37^. We chose to use Exenatide as the intervention since had approval for intravenous use and because similar doses and administration had been applied in previous trials and seemed to be safe and efficacious^18,35^. The infusion had a significant effect on perioperative blood sugar concentrations. However, we cannot rule out that a different regimen for administration or longer-term use of Exenatide may have had other effects.

## Conclusion

In conclusion, a perioperative infusion with the GLP-1 analog exenatide did not reduce mortality or morbidity from stroke, renal failure, or heart failure in patients undergoing heart surgery with cardiopulmonary bypass. There were no differences in safety endpoints.

## Data Availability

Data can be shared upon request

## Acknowledgements

The trial steering committee would like to extent our sincere gratitude to the clinical staff at Department of Cardiothoracic Anesthesiology and Intensive care including the post-surgical fast-track unit, the clinical staff at Department of Cardiothoracic Surgery, and the clinical staff at Cardiac Intensive Care Unit 2143. Your dedication remains crucial for the conduct of clinical trials in the Heart Centre. Furthermore, we would like to thank Clinical Research Unit (Klinisk Forskningsenhed) for their aid in day-to-day conduct of the trial.

## Funding statement

This work was supported by:

Læge Sofus Carl Emil Friis og Hustru Olga Doris Friis’ Legat. 2017 (DKK 461,180)

Aase og Ejnar Danielsens Fond 2017, grant no. 10-001976 (DKK 200,000)

Danish Heart Foundation 2019, Grant number 19-R133-A9174-22144 (DKK 1,000,000)

The Heart Centre Research Foundation 2017, (DKK 160,000)

Lundbeck Foundation (R186-2015-2132)

